# Importance of E484K and N501Y mutations in SARS-CoV-2 for genomic surveillance: rapid detection by restriction enzyme analysis

**DOI:** 10.1101/2021.05.04.21256650

**Authors:** Rossana C Jaspe, Yoneira Sulbaran, Carmen L Loureiro, Pierina D’Angelo, Lieska Rodríguez, Domingo J Garzaro, Héctor R Rangel, Flor H Pujol

**Affiliations:** Laboratorio de Virología Molecular, Centro de Microbiología y Biología Celular, Instituto Venezolano de Investigaciones Científicas, Caracas, Miranda, Venezuela; Instituto Nacional de Higiene “Rafael Rangel”, Caracas, Miranda, Venezuela

**Keywords:** COVID-19, SARS-CoV-2, Variants of Concern (VOC), RFLP, Rapid screening, Mutations

## Abstract

**Introduction:** Variants of Concern of SARS-CoV-2 (VOCs), the new coronavirus responsible for COVID-19, have emerged in several countries. Two mutations in the gene coding for the Spike protein of the viral genome are particularly important and associated with some of these variants: E484K and N501Y. Restriction enzyme analysis is proposed as a rapid method to detect these two mutations.

**Methodology:** A search on GISAID was performed in April 2021 to detect the frequency of these two mutations in the sequence available and their association with other lineages. A small amplicon from the Spike gene was digested with two enzymes: HpyAV, which allows detecting E484K mutation, and MseI, for detecting the N501Y one.

**Results:** The mutations E484K and N501Y, associated with VOCs, have emerged in several other lineages, particularly E484K. A 100% correlation was observed with sequencing results.

**Conclusions:** The proposed methodology, which allows screening a great number of samples, will probably help to provide more information on the prevalence and epidemiology of these mutations worldwide, to select the candidates for whole-genome sequencing.

## Introduction

In March 2020, the COVID-19 pandemic was declared, caused by an emerging coronavirus, SARS-CoV-2. One year later, this infection has caused more than 100 million cases and more than 2 million deaths worldwide. This virus belongs to the family *Coronaviridae*, order *Nidovirales*. This viral order is unique among the RNA viruses since the viruses belonging to this order encode for an exonuclease, which enables proof-reading capacity to the replication machinery, limiting mutational events. However, the tremendous number of replication events that this virus has undergone, in addition to an elevated frequency of recombination, and to the probable action of host deaminases on the viral genome [1], has allowed the emergence of many point mutation and frequent deletions in the viral genome [2].

Different variants (groups of viruses sharing particular types of mutations) have emerged at the end of 2020. Some of these variants have been defined of Interest (VOI) or Concern (VOC) by WHO: the variant B1.1.7 (VOC), for which an increase in incidence after its emergence was observed in the UK, variant B.1.351 (VOC), with an increase in prevalence in South Africa, and variants B.1.1.28.1 or P.1 (VOC) and B.1.1.28.2 or P.2 (VOI), which are now predominating in Brazil, are examples of these variants (variants named according to Pango lineages classification (https://cov-lineages.org/lineages.html) [3-5].

Some of the mutations harbored by these variants are of public health concern for two main reasons: mutation N501Y might lead to increased transmissibility of the viruses harboring this mutation, and mutation E484K, has been frequently associated with reinfection cases, and might reduce the neutralizing activity of antibodies produced by vaccination [6,7]. Genomic surveillance has been proposed for monitoring the introduction of SARS-CoV-2 Variants of Concern (VOCs) in different countries [8].

A rapid method for detecting those variants might be useful for screening large quantities of samples, in settings where sequencing facilities are limited. This would be very useful for rapid and massive screening of these variants. In this study, we propose a simple restriction enzyme digestion analysis for the detection of two key mutations in the SARS-CoV-2 Spike: E484K and N501Y.

## Methodology

### Analysis of sequences available at GISAID for E484K and N501Y mutations

Sequences available at GISAID on April 24 were analyzed for the presence of E484K and N501Y, at https://www.gisaid.org/phylodynamics/global/nextstrain/ and https://www.epicov.org/.

### RT-PCR

This study was approved by the Bioethical Committee of IVIC. We previously detected in Venezuela, the circulation of variants of the B.1.1.28.1 lineage (P1-like) and samples harboring the E484K mutation without the N501Y one, by sequencing part of the Spike gene in Venezuelan isolates (Jaspe, RC, in preparation). RNA from clinical samples positive by qRT-PCR (classified as wild-type, WT, or harboring E484K alone or with N501Y) was amplified with primers 76.1L (5’-CCAGATGATTTTACAGGCTGCG-3’) and 76.8R (5’-GTTGCTGGTGCATGTAGAAGTTC-3’) using SuperScript III One-Step RT-PCR System with Platinum Taq High Fidelity DNA Polymerase (Thermo Fisher Scientific), and the following PCR conditions: an incubation at 55°C for 30 min, followed by 94°C/3min and 40 cycles of 94°C/15 sec, 55°C/30 sec and 68°C/30 sec, with a final extension of 68°C for 7 min. Superscript II One-Step PCR System with Platinum Taq DNA Polymerase (Thermo Fisher Scientific), was also used successfully.

### Restriction analysis

Five µl of the amplicon were digested with 1 unit of HpyAV or MseI for 1 hour at 37°C and then loaded in a 3% agarose gel electrophoresis for band visualization with Ethidium bromide. Restriction results were compared with the sequence obtained by sending PCR purified fragments to Macrogen Sequencing Service (Macrogen, Korea).

## Results

A total of 1,224,815 sequences of SARS-CoV-2 were available on April 24 at GISAID. Figure 1 shows the frequency of sequences harboring the N501Y, E484K, or both mutations. However, this frequency does not reflect the true prevalence of these mutations worldwide, since a strong bias exists, which has been accentuating over the last months. There are strong differences in genomic sequencing capacities between developed and developing countries, and even between developed countries (Table 1).

**Table 1.**
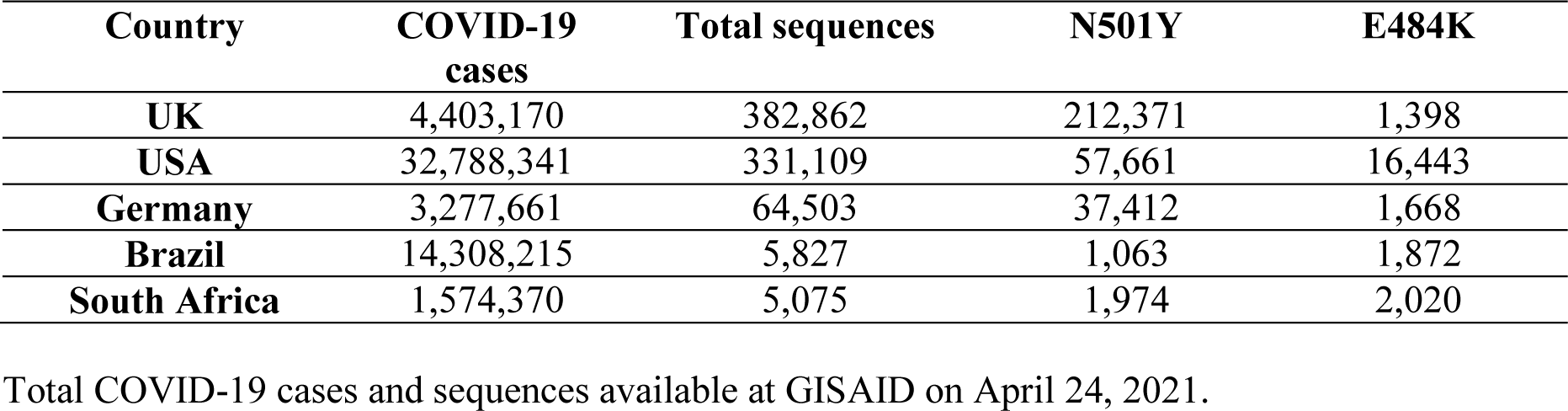
Number of sequences harboring E484K or N501Y mutations in selected countries

**Figure 1.**
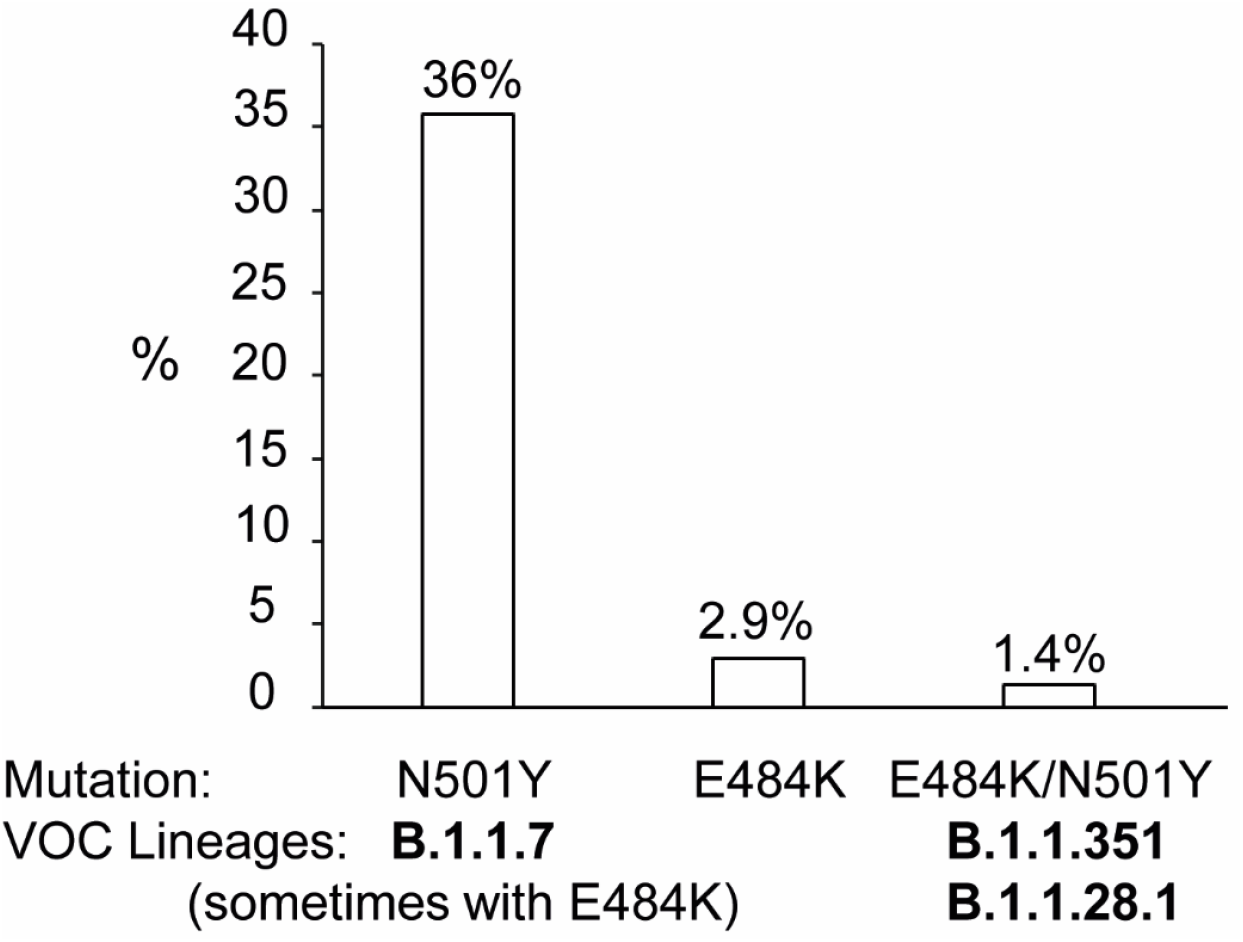
Frequency of sequences in GISAID harboring N501Y, E484K mutations or both.

For example, UK is the country that has provided the highest number of sequences to GISAID, more than the USA, even if the total number of COVID-19 cases is 7.4 times lower than the ones in the USA. VOC of the lineage B.1.1.7, which lacks the E484K mutations, emerged in the UK and is also frequent in the USA. In contrast, the other two VOCs, which harbor E484K in addition to N501Y, emerged in Brazil and South Africa, countries with robust sequencing capacities but not comparable to the countries previously mentioned (Table 1). This is the main reason for the higher abundancy of N501Y mutation, compared to E484K one (Figure 1).

Even if the frequency of the E484K mutation appears as relatively low in the sequences available at GISAID, this mutation has been found in some isolates from several lineages, more abundant than the ones where N501Y mutation has been described, in spite of the significant lower number of sequences available in GISAID, harboring this mutation (Table 2).

**Table 2.** Lineages where E484K or N501Y mutations can be found

| Mutation | Lineages where some or all sequences harbor this mutation |
| --- | --- |
| **E484K** | **Total lineages: 127** |
| Lineages with less than 10 sequences with the mutation (n=99) | A A.23 B B.1.1.1 B.1.1.10 B.1.1.101 B.1.1.115 B.1.1.16 B.1.1.161 B.1.1.214 B.1.1.216 B.1.1.220 B.1.1.241 B.1.1.25 B.1.1.270 B.1.1.273 B.1.1.297 B.1.1.316 B.1.1.317 B.1.1.322 B.1.1.34 B.1.1.348 B.1.1.351 B.1.1.365 B.1.1.38 B.1.1.393 B.1.1.398 B.1.1.406 B.1.1.412 B.1.1.434 B.1.1.461 B.1.1.482 B.1.1.486 B.1.1.51 B.1.1.515 B.1.1.519 B.1.1.57 B.1.110 B.1.111 B.1.131 B.1.134 B.1.139 B.1.160.26 B.1.177.10 B.1.177.12 B.1.177.31 B.1.177.4 B.1.177.40 B.1.177.44 B.1.177.51 B.1.177.62 B.1.177.77 B.1.177.85 B.1.177.86 B.1.214.3 B.1.218 B.1.220 B.1.221 B.1.234 B.1.237 B.1.241 B.1.243 B.1.258 B.1.265 B.1.279 B.1.281 B.1.311 B.1.313 B.1.314 B.1.349 B.1.36 B.1.36.10 B.1.361 B.1.369 B.1.384 B.1.395 B.1.396 B.1.409 B.1.411 B.1.416 B.1.429 B.1.441 B.1.486 B.1.526.1 B.1.526.2 B.1.527 B.1.538 B.1.555 B.1.564 B.1.573 B.1.577 B.1.595 B.1.609 B.1.612 B.1.620 C.11 C.22 L.3 P.1.1 |
| Lineages with 10-100 sequences with the mutation (n=18) | A.23.1 B.1.1 B.1.1.28 B.1.1.306 B.1.1.318 B.1.1.345 B.1.110.3 B.1.160 B.1.177 B.1.177.88 B.1.243.1 B.1.375 B.1.400 B.1.596 B.1.618 N.9 P.3 R.2 |
| Lineages with more than 100 sequences with the mutation (n=10) | B.1.2 (104) B.1.619 (167) B.1.1.207 (190) **B.1.1.7\*** (378) B.1 (403) P.2 (1.896) R.1 (2.175) **P.1** (4.192) B.1.526 (6.011) **B.1.351** (7.595) |
| **N501Y** | **Total lineages: 111** |
| Lineages with less than 10 sequences with the mutation (n=87) | A.2.52 A.21 A.23 A.25 AS.2 B B.1.1.10 B.1.1.28 B.1.1.34 B.1.1.368 B.1.1.372 B.1.1.375 B.1.1.378 B.1.1.419 B.1.1.420 B.1.1.507 B.1.1.519 B.1.1.521 B.1.104 B.1.110.3 B.1.149 B.1.160 B.1.160.7 B.1.165 B.1.177.31 B.1.177.32 B.1.177.35 B.1.177.4 B.1.177.44 B.1.177.50 B.1.177.51 B.1.177.73 B.1.177.81 B.1.177.82 B.1.177.86 B.1.218 B.1.221 B.1.258 B.1.258.17 B.1.289 B.1.324 B.1.36 B.1.36.1 B.1.36.10 B.1.36.7 B.1.367 B.1.37 B.1.375 B.1.395 B.1.404 B.1.408 B.1.427 B.1.429 B.1.459 B.1.470 B.1.474 B.1.486 B.1.495 B.1.525 B.1.526 B.1.526.2 B.1.527 B.1.530 B.1.533 B.1.577 B.1.595 B1.1.1 B1.1.122 B1.1.134 B1.1.161 B1.1.166 B1.1.216 B1.1.220 B1.1.231 B1.1.302 B1.1.312 B1.1.317 B1.1.322 B1.1.333 B1.1.335 B1.1.355 B1.1.38 B1.1.50 B1.1.58 B1.1.99 C.8 P.1.1 |
| Lineages with 10-100 sequences with the mutation (n=15) | A B.1.126 B.1.153 B.1.177 B.1.243 B.1.311 B.1.36.31 B.1.480 B.1.596 B.1.596.1 B.1.597 B1.1.136 B1.1.306 C.11 P.3 |
| Lineages with more than 100 sequences with the mutation (n=9) | A.27 (207) AP.1 (554) B.1 (777) B.1.1 (238) **B.1.1.7** (342.782) B.1.2 (713) **B.1.351** (7.613) B1.1.189 (158) **P.1** (4.195) |

\*Some isolates belonging to the B.1.1.7 have acquired the E484K mutation.

Figure 2 shows the restriction pattern of Wild Type (WT) samples and isolates harboring mutations E484K and N501Y (M). Digestion with HpyAV of the 293 bp amplicon of the Spike partial genomic region yielded two bands of 173 and 120 bp for samples with E484, while the digestion site is abolished with K484. Digestion with MseI yielded a fragment of 182 and 111 for samples with N501, and a fragment of 173, 111 and 9 bp (nor observed in the gel) for samples with Y501 (Figure 2A). The 9 bp difference of the product between the WT and Y501 mutated samples was easily differentiated in 3% agarose gels (Figure 2B). NuSieve agarose or Polyacrylamide gel electrophoresis can also be used for more discrimination, particularly of bands of 173 and 182 bp. However, this was not necessary in our hands.

**Figure 2.**
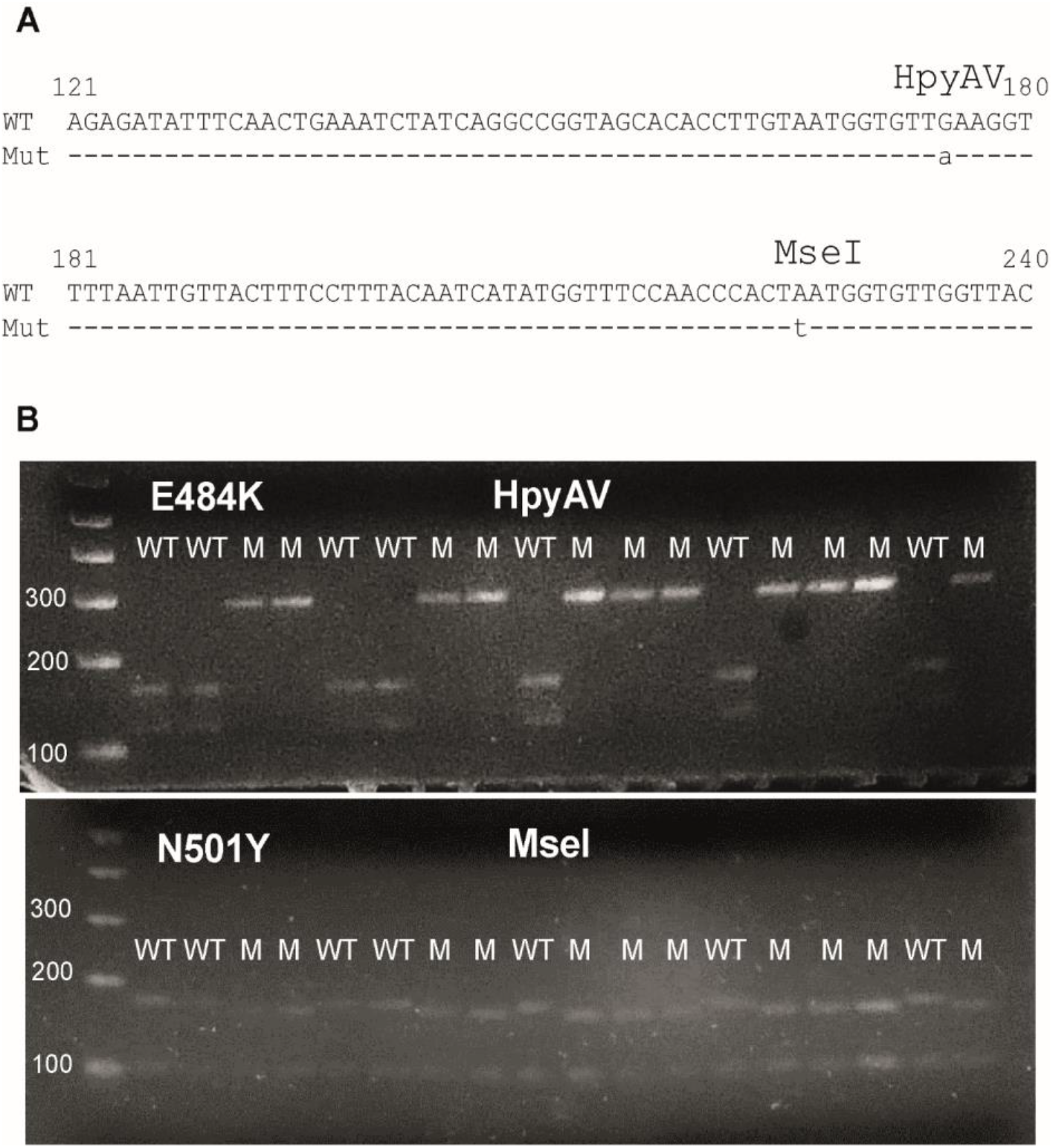
Restriction analysis of amplicons with E484K or N501Y mutations. A. Sequence of the amplified product showing the restriction sites which discriminate Wild-type (WT) or mutant (Mut or M) viruses. B. Agarose gel electrophoresis of digested PCR-amplified products. PCR-amplified products digested with the enzyme shown were run with molecular weight markers (1Kb plus DNA ladder): smaller bands are signaled (100, 200 and 300 bp).

A total of 40 samples (12 WT, 27 with both mutations and 1 with only E484K) were analyzed for their restriction pattern and compared to the presence or not of the two mutations in their sequence. A 100% correlation was observed in the detection of the mutations between the two methods (data not shown).

## Discussion

SARS-CoV-2 variants are emerging and spreading rapidly in several parts of the world [3]. Mutations E484K and N501Y have been recurrently associated with many of the VOCs and may appear in other variants already unknown. Indeed, a survey of sequences available at GIASID showed that these mutations, particularly E484K, have been found in sequences for many lineages. As stated before, the E484K mutation has been found in cases of reinfection and may represent a mutation that emerged to escape the presence of neutralizing antibodies [9]. The frequency of variants or mutations cannot be estimated with the sequences available at GISAID, since huge disparities exist between the sequencing capacities among countries.

The presence of any of these two mutations does not necessarily represent that a VOC is circulating in a specific country. Their presence does not necessarily mean that this isolate will gain the phenotypic advantages provided by these mutations in VOCs, as the increased transmissibility observed for the VOCs and provided by N501Y. Many other mutations present in these VOCs (point mutations and deletions) might be contributing to the enhanced transmissibility of these VOCs [10].

However, the specific detection of these two mutations, which play a key role in determining the phenotype of the isolate, might be of particular interest.

Thus a rapid method for identifying those mutations should be very useful, particularly in settings where massive whole genome sequencing is not available. Rapid methodologies might be used for the rapid screening of several samples. The whole protocol can be run in a day.

## Conclusions

The proposed methodology allows analyzing a great number of samples to select the ones that may harbor mutations of concern, before proceeding to whole genome sequencing. On the other hand, once the presence of the variants is confirmed by whole genome sequencing, this method can be used for the rapid estimation of their prevalence in different geographical regions.

## Data Availability

All data is available.

## Acknowledgements

This study was supported by Ministerio del Poder Popular de Ciencia, Tecnología e Innovación of Venezuela.

## Notes

### Competing Interest Statement

The authors have declared no competing interest.

### Funding Statement

This study was supported by Ministerio del Poder Popular de Ciencia, Tecnologia e Innovacion of Venezuela.

### Author Declarations

This study was approved by the Bioethical Committee of IVIC.

